# Cardiac Troponin T is a Serum Biomarker of Respiratory Dysfunction in Amyotrophic Lateral Sclerosis

**DOI:** 10.1101/2023.12.31.23300684

**Authors:** Teresa Koch, Rachel Fabian, Leonie Weinhold, Franz-W. Koch, Saman Barakat, Sergio Castro-Gomez, Torsten Grehl, Sarah Bernsen, Patrick Weydt

## Abstract

**Objective:** Informative biomarkers are an urgent need in management and therapy development of amyotrophic lateral sclerosis. Serum cardiac troponin T is elevated in most amyotrophic lateral sclerosis patients and not correlated with neurofilaments. We sought to delineate the functional implications and the informative value of serum troponin T with regard to respiratory function, a major prognostic factor in amyotrophic lateral sclerosis.

**Methods:** We analyzed two independent hospital-based amyotrophic lateral sclerosis cohorts (d=discovery cohort; v= validation cohort) with data available on serum cardiac troponin T levels (n_d_=297; n_v_=49), serum neurofilament light chain levels (n_d_=116; n_v_=17), and routine respiratory test results (n_d_=86; n_v_=49).

**Results:** Serum cardiac troponin T levels, unlike serum neurofilaments, were strongly associated with the respiratory domain of the revised amyotrophic lateral sclerosis functional rating score (r_d_ = - 0.29, p_d_ = 0.001; r_v_= - 0.48, p_v_ = 0.007) and with relevant pulmonary function parameters (n_d_), namely SVC% (r = - 0.45; p = 0.001), FVC% (r = - 0.43; p = 0.001), FEV1% (r = −0.37, p = 0.007), and PEF (r = - 0.34, p = 0.027).

Serum cardiac Troponin T reliably discriminated benchmarks of SVC% < 80%: (AUC 0.75, p = 0.003), FVC % < 80%: (AUC 0.72, p = 0.011) and PEF% <75%: (AUC 0.72, p = 0.015).

**Interpretation:** Our findings confirm cardiac Troponin T as an informative serum biomarker in amyotrophic lateral sclerosis, complementing neurofilaments. Serum Troponin T can flag compromised respiratory function in amyotrophic lateral sclerosis and might prove useful as a proxy of respiratory impairment with prognostic implications.

## Introduction

Amyotrophic lateral sclerosis (ALS) is a fatal neurodegenerative disorder characterized by the progressive degeneration of central and peripheral motor neurons. This leads to skeletal muscle weakness and wasting, typically culminating in respiratory failure within very few years^1^. Currently, treatment options for ALS are limited. Therapy development and the challenging disease management require reliable biomarkers that can guide treatment strategies and help define critical disease stages and prognosis^2^. Neurofilament heavy and light chain (NfL) reflect neuroaxonal damage and are readily quantifiable in blood and cerebrospinal fluid. They aid the diagnosis, prognosis, and have been accepted as therapy response markers by regulatory authorities^3–5^.

Serum cardiac Troponin T (cTnT), widely in use as a serum marker of myocardial injury, has recently emerged as a promising candidate serum biomarker for ALS, correlating with disease severity and progression^6,7^. Accumulating evidence suggests that degenerating or regenerating skeletal muscle is an extracardiac source of serum cTnT, and can lead to serum cTnT elevations independent of myocardial damage^8–10^.

Respiratory function is one of the main determinants of survival in ALS. Respiratory impairment may occur at any time during the highly variable course of the disease, and its clinical symptoms initially manifest subtly and unspecifically^11,12^. Early recognition of respiratory dysfunction helps to ensure that non-invasive ventilation (NIV) can be discussed in advance, one of the most life-prolonging therapies available^13,14^. Moreover, the quantification of the respiratory impairment helps gauge individual prognosis^15^.Various tests are employed to determine respiratory impairment, but differ in complexity and relevance ^16^

The respiratory domain of the ALSFRSr, while well established as a predictor of survival^17^, is a comparatively crude three item, 12-point measure of respiratory function. In a clinical setting respiratory function is commonly evaluated with a range of lung parameters that indirectly capture the inspiratory and expiratory muscle functions in pulmonary function testing (PFT).

Among these test modalities slow vital capacity (SVC) and forced vital capacity (FVC) of PFT are the most widely used. In ALS, they are recommended to monitor respiratory impairment and provide clinical triggers for initiating interventions like NIV and percutaneous gastrostomy placement^18–20^. The objective of the present study was to investigate the relationship between cTnT serum levels and respiratory dysfunction as captured by ALSFRr and PFT.

## Methods

### Study Design and Patient Characteristics

The present study was conducted on two independent hospital-based cohorts, one at the University Hospital Bonn and one at the Krupp Hospital Essen (Figure 1). We first reviewed the files of all patients seen in the ALS clinic of University Hospital Bonn from 01 of March 2019 to 31 of December 2021 (discovery cohort, n_d_=297). We included patients with a diagnosis of ALS according to the revised El Escorial Criteria documented in the clinical files by board-certified neurologists (PW; RF; SB). In all patients at the Bonn clinic, ALS functioning rating score revised (ALSFRSr) and serum cTnT are collected as part of the clinical routine. For 96 of these patients PFT measurements (SVC, FVC, PEF, FEV1, IC, TLC) from a time window of +/- 31 days of the serum sampling were available.

**FIGURE 1:**
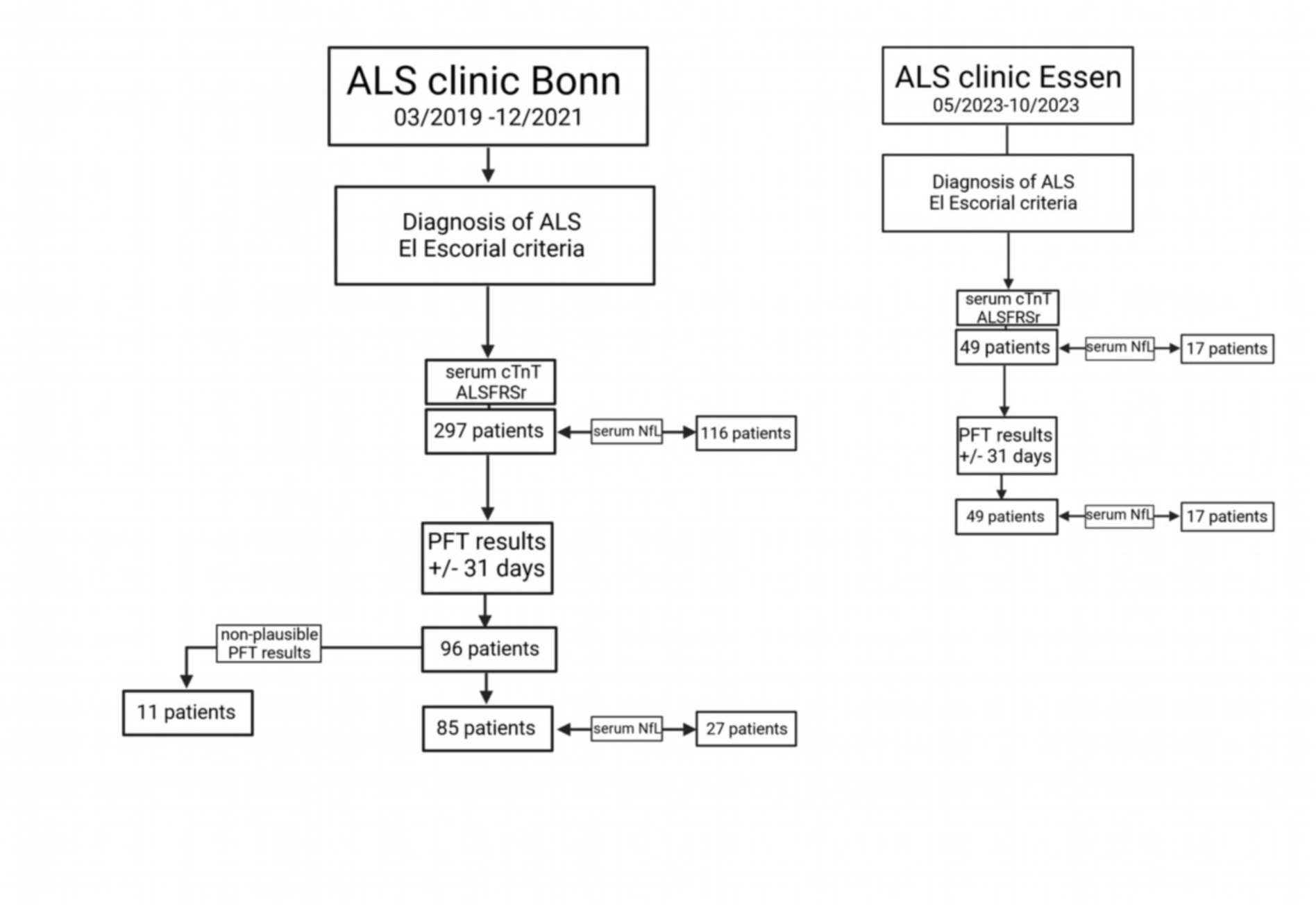
Flow chart of retrospective study. The retrospective analysis of two independent hospital cohorts (discovery cohort Hospital Bonn, 2019-2021; validation cohort Hospital Essen, 2023) was performed targeting the same clinical characteristics: the diagnosis of ALS following the El Escorial criteria, serum cTnT, serum NfL measurements, and ALSFRSr scale. Results of pulmonary function tests were included when available +/-31days to the latter. ALS = Amyotrophic lateral sclerosis; ALSFRSr = revised ALS functional rating; cTnT = cardiac Troponin T; PFT = pulmonary function test; serum NfL = serum Neurofilament light chain.

Eleven of these PFT measurements were eliminated for lack of plausibility after consultation with a board-certified pulmonologist (FWK).

Serum NfL testing was implemented in routine testing with a lag of several months and thus is only available from a subset of the Bonn patients (n_d_ =116). Demographic data included sex, age, height, weight, and body mass index (BMI). Disease history data included date and site of ALS symptom onset, region of ALS onset and disease duration.

The Essen cohort served as an independent validation cohort (n_v_=49) with available data on ALSFRSr, serum cTnT, serum NfL, and PFT (SVC).

As the laboratory measurements and clinical assessments were part of the routine clinical work up andretrospectively analyzed, no formal consent was needed per statement of our institutional ethics review board (Ethics Board decision letter 324/20).

### Biomarker measurements

Serum high-sensitivity cTnT was measured using electrochemiluminescence immunoassay (ECLIA) in a commercial laboratory (Labor Volkmann Karlsruhe, Germany) as described previously^6^. Serum NfL was measured using commercially available enzyme-linked immunosorbent assay kits for NF-L (IBL, Hamburg, Germany), also as described previously^21^.

### Respiratory function assessments

PFT readouts were all obtained with a Vyntus® Body Plethysmograph (Vyaire Medical GmbH Höchberg) at the Bonn University Pulmonology department. In the Essen clinic SVC measurements are part of the routine work up and obtained with a Vitalograph Spitotac MODEL 7000 (Vitalograph GmbH Hamburg).

### Statistical analysis

Patient characteristics are presented as absolute and relative frequencies for categorical variables and as means with standard deviations (SDs) or median (range) for continuous variables. For analysis, the biomarker serum cTnT and serum NfL were log-transformed (natural logarithm).

To assess the association between cTnT with pulmonary function tests and ALSFRSr, Pearson correlation coefficients were calculated. Correlation coefficients of serum cTnT and serum NfL were compared using the Fisher z test, according to^22,23^. Additionally, simple and multiple linear regression analyses with cTnT as independent variable and SVC%, FVC%, FEV1%, PEF%, IC% and TLC% as outcome variables were performed. Multiple linear regression analyses were adjusted for sex, site of onset BMI and age. Addressing sex differences in this rare disease with a small sex bias was beyond the scope of this manuscript.

To calculate the diagnostic utility of cTnT and NfL for the distinction of clinically relevant thresholds of SVC, FVC (80% and 50% respectively) and PEF (75%, 50%), the area under the curve (AUC) was calculated with receiver operating characteristic (ROC) curve analyses with confidence intervals. The optimal cut-off points we determined based on the Youden Index.

All analyses were initially conducted using the discovery cohort and significant findings were subsequently tested in the validation cohort.

The significance level was set to 0.05 and p-values were adjusted by the Bonferroni correction for multiple testing.

All statistical analyses were performed with GraphPad Prism® (10) (GraphPad Software Inc., San Diego, USA). Microsoft Excel 2019 ® was used for data storage (Microsoft Redmond, Washington, USA). Biorender.com was utilized for figures BioRender (Toronto, ON Canada).

### Data availability statement

Anonymized data will be made available upon reasonable requests to the corresponding author. The data are not publicly available due to privacy or ethical restrictions.

## Results

### Clinical characteristics

The basic characteristics of the discovery cohort and the validation cohort are described in Table 1. The two cohorts were similar regarding key features such as age, sex ratio, ALSFRSr score and proportion of cases with spinal vs. bulbar onset. The discovery cohort (Bonn) had a significantly longer mean disease duration at sample collection (29.7 months) than the validation cohort (Essen) (6.4 months), reflecting the earlier initiation of the study. The mean of serum cTnT levels in the discovery cohort was 30.4 ng/l (± 38.6), i.e. >2 times above the upper reference limit (URL) of 14.0 ng/l. 192 (65.0 %) of 297 patients had elevated serum cTnT. In the validation cohort 24 of measurements (47.0%) were elevated with a similar mean of 27.4 ng/l (±42.3). This is in line with previous reports^7,24^. The mean serum NfL was 110.5 pg/ml(±87.3) and elevated in 94 patients (81.0%) of 116 patients in the discovery cohort. Similarly, the mean NfL serum level of the validation cohort was elevated at 129.5 pg/ml (±58.0), albeit in a smaller proportion of cases (41.2 %).

**TABLE 1:**
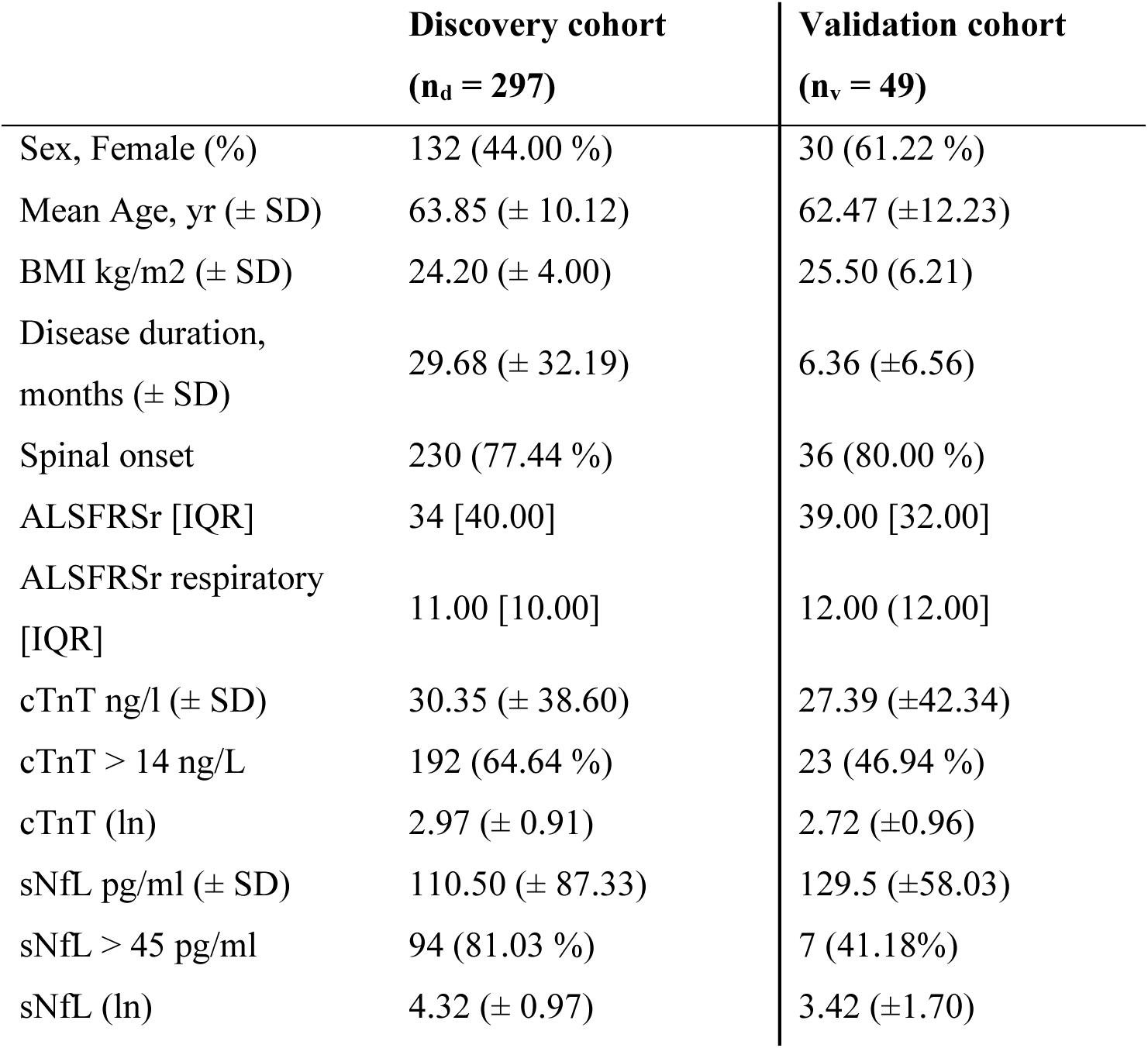
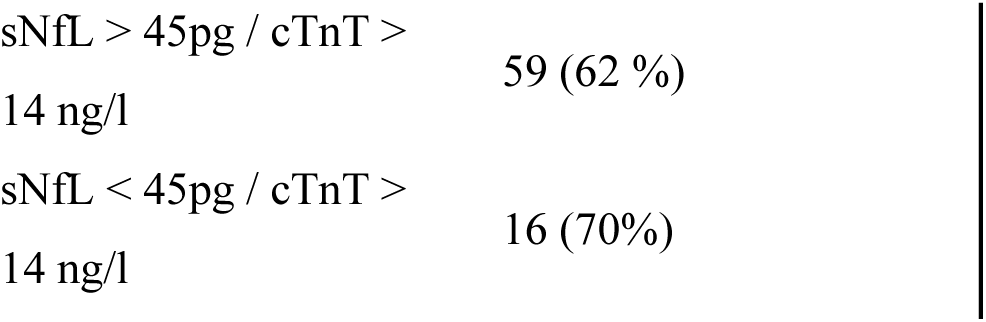
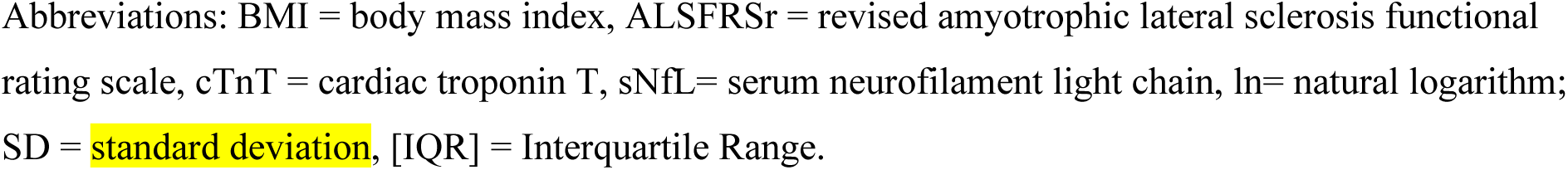
Baseline characteristics of the discovery cohort and the validation cohort.

### Correlations with clinical parameters and pulmonary function

Serum cTnT levels correlated strongly and negatively with the total ALSFRSr (r = - 0.32, p = 0.001), and – pertinent to the present study – with its respiratory domain (r= - 0.29, p = 0.001) (Figure 2). This correlation was fully recapitulated in the validation cohort (r =-0.40, p = 0.055). The serum NfL levels, in sharp contrast, while correlating with the total ALSFRSr, showed no significant correlation with the respiratory domain. As respiratory function is one of the most important prognostic factors in ALS^15,25^ we were compelled to further probe the relationship of serum cTnT with respiration. We found that serum cTnT showed significant negative associations with four out of six selected parameters, namely: FVC (− 0.43, p = 0.001), SVC (− 0.45, p = 0.001), FEV1(− 0.37, p = 0.007), PEF (−0.34, p = 0.027) (Figure 3). The associations with SVC were confirmed in the validation cohort (data not shown).

**FIGURE 2:**
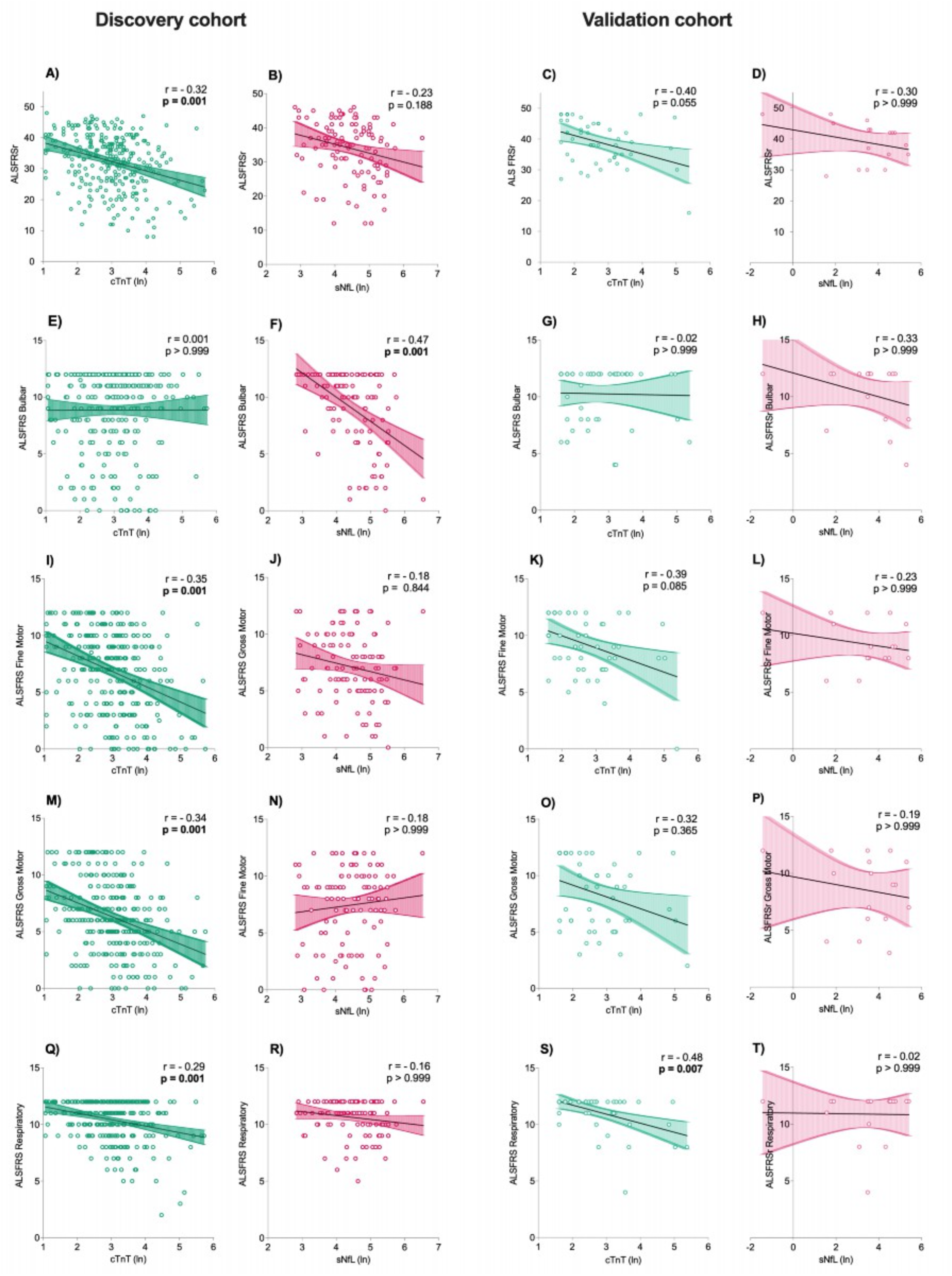
Correlation plots of serum cTnT and sNfL with ALSFRSr and its respiratory subcategory. All four ALSFRS domains (bulbar, gross motor, fine motor, respiratory) contain three items. (A-H) Correlation and linear regression analyses were performed with the natural logarithm(ln) of cTnT and sNfL using parametric Pearson correlation coefficient r. Colored areas represent 95% CI. Curves demonstrate correlations in the discovery cohort between serum cTnT with ALSFRSr (A), and the four domains (E, I, M, Q) compared with correlations between sNfL and ALSFRSr (B) and the four domains (F, J, N, R) and in the validation cohort between serum cTnT with ALSFRSr (C) and the four domains (G, K, O, S) compared to correlations of sNfL with ALSFRSr (D), and with the four domains (H, L, P, T). Abbreviations: ALSFRSr = revised ALS functional rating scale; cTnT = cardiac Troponin T; sNfL = serum Neurofilament light chain.cTnT=cardiac Troponin T; sNfL=serum Neurofilament light chain; ln = natural logarithm, r= Pearson correlation coefficient

**FIGURE 3:**
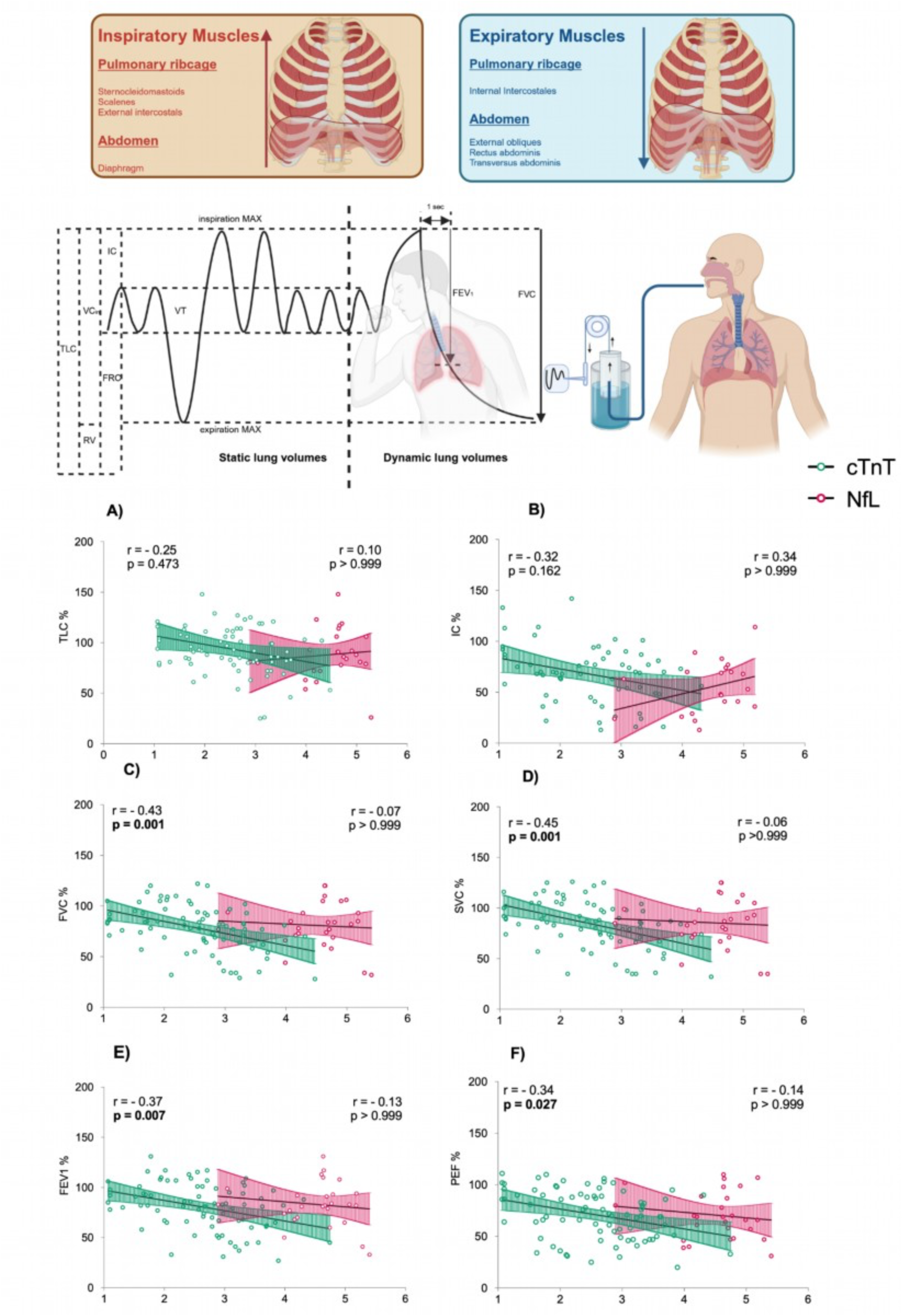
Correlation of serum cTnT and serum NfL with parameters of pulmonary function testing. Upper Figure: the schematic panel shows inspiratory and expiratory muscle groups and the principle of pulmonary function testing (PFT). Below: Correlation plots between natural logarithm of serum cardiac Troponin T (TnT ln) and serum Neurofilament light chain (NfL ln) and (A) TLC% = Total lung capacity, (B) IC% = of static lung volumes; (C) FVC% = forced vital capacity, (D) SVC% = slow vital capacity; (E) FEV1% = Forced expiration in first second (F) PEF% = peak expiratory flow. Correlation and linear regression analyses were performed using parametric Pearson correlation coefficient r. Colored areas represent 95% CI.

The association between serum cTnT and the PFT parameters was further confirmed using multiple linear regression analysis after adjusting for the confounding factors sex, subtype, BMI and age. Serum cTnT was most significantly associated with FVC (p *=* 0.001, CI = −19.8 to −6.9) SVC (p *=* 0.003, CI = −19.4 to −6.3) and FEV1 (p *=* 0.003, CI = −17.9 to −5.9) (Figure 4).

**FIGURE 4:**
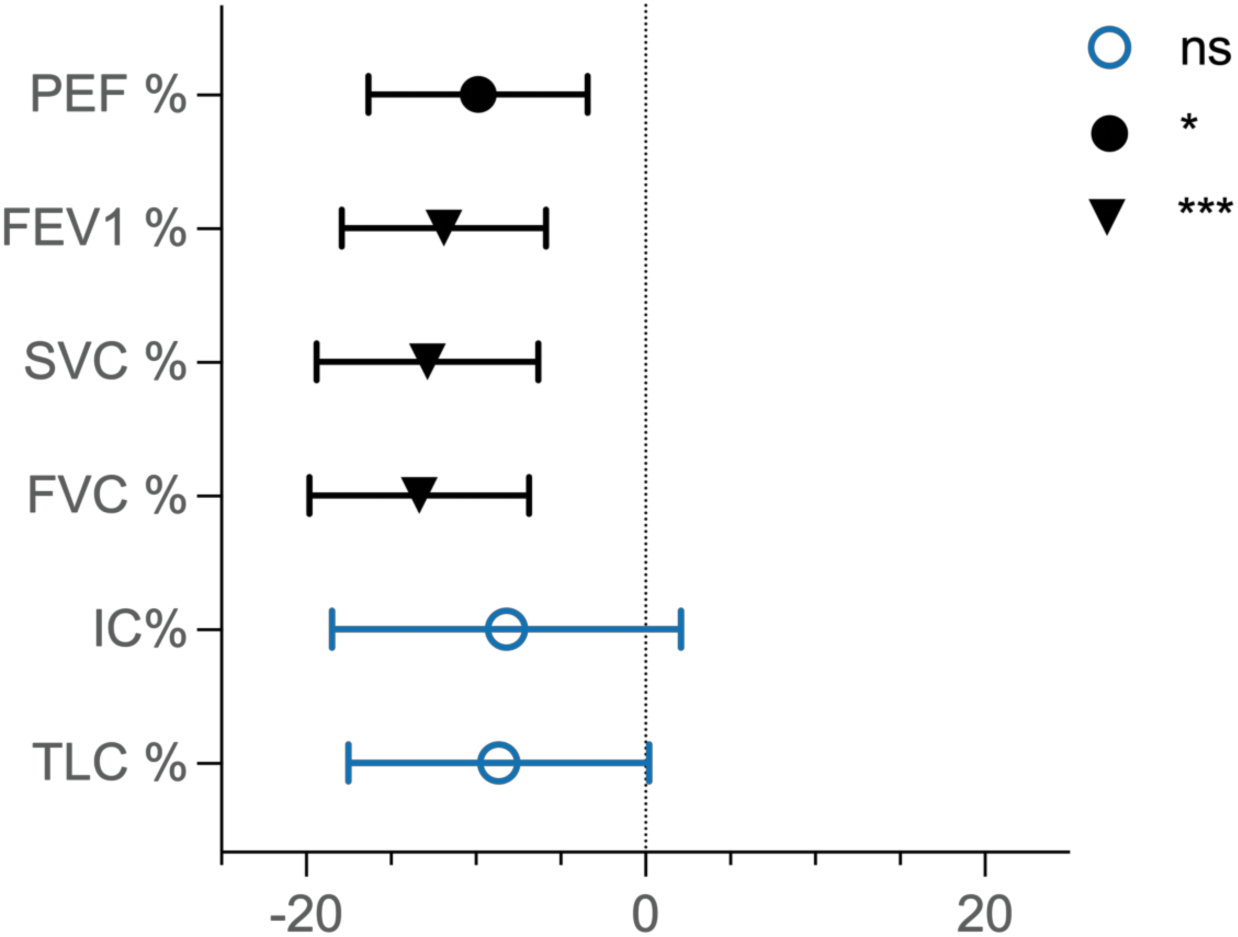
Forest plot of PFT parameters estimates for serum cTnT as independent variable. Estimates of PFT parameters as the independent variable (natural logarithm of serum cTnT) changes. Estimates parameters are graphed with CI as error bar and p values as variating symbols: for TLC = - 8.7, (CI = −17.5 to 0.2, p = 0.776), for IC % = −8.2, (CI = −18.5 to - 2.1, p > 0.999), for FVC% = −13.3 (CI =-19.8 to - 6.9, p = 0.001), for SVC% = −12.9 (CI = - 19.4 to - 6.3, p =0.003), for FEV1% = −11.9. (CI = −17.9 to - 5.9, p = 0.003) and for PEF% = −9.9 (CI =- 16.3 to −3.4, p =0.042). Abbreviations: TLC% = Total lung capacity; IC%= Inspiratory capacity; FVC%= Forced Vital Capacity; VC%=Vital Capacity; FEV1%= Forced Expiratory Volume in the first second; PEF%=Peak Expiratory Flow; *=p- value, ns-= non-significant, CI = confidence interval

To test the diagnostic potential of serum cTnT levels to flag PFT declines below clinically important thresholds, we generated ROC curves (Figure 5).

**FIGURE 5:**
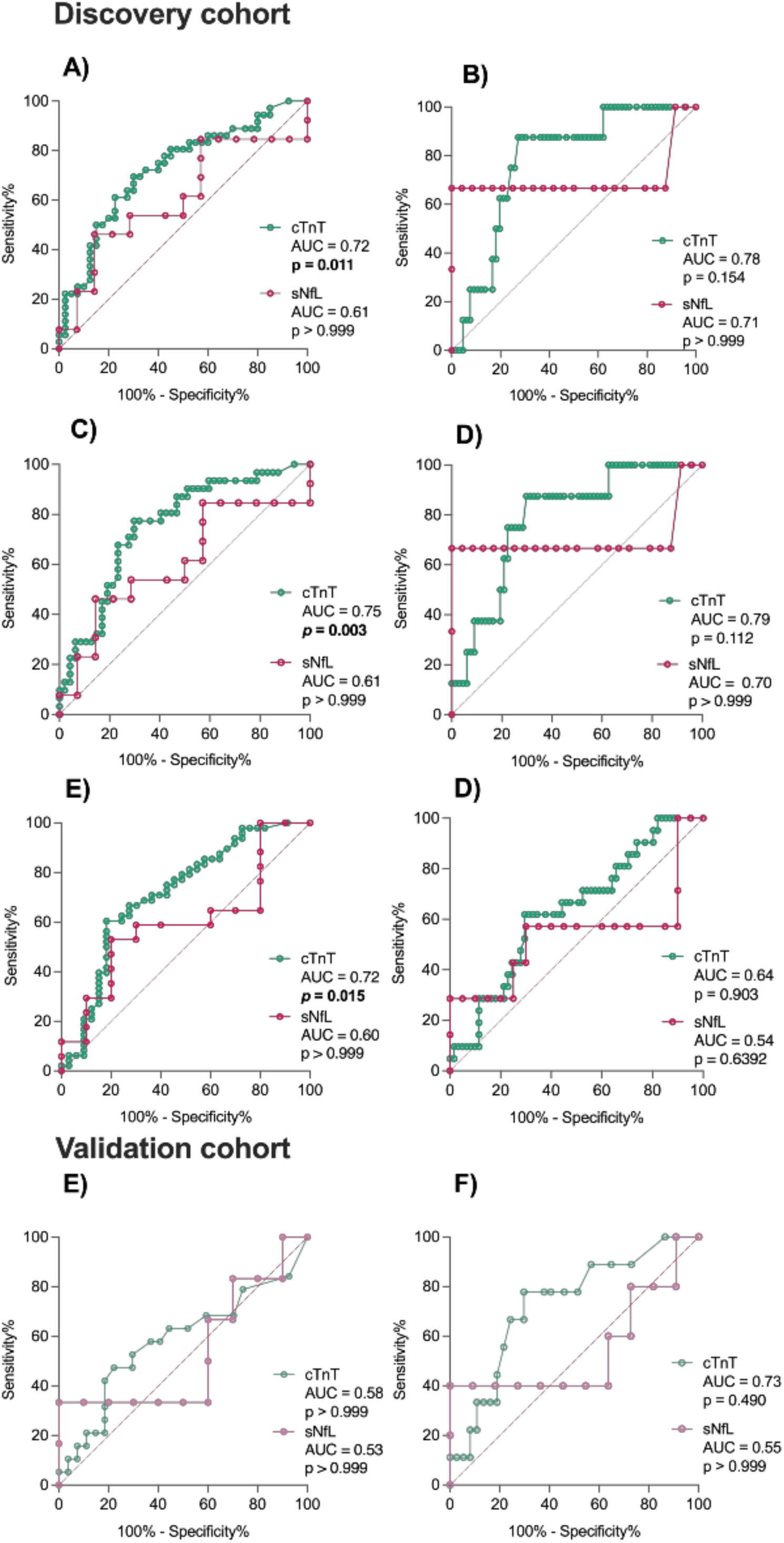
The diagnostic potential of cTnT versus NfL to distinguish benchmarks of pulmonary function. Receiver Operating Characteristics (ROC) Curvesanalyze the diagnostic accuracy of serum cTnT and serum NfL distinguishing clinically relevant thresholds of PFT parameters. (A) FVC < 80%, (B) FVC < 50%, (C) SVC < 80 %, (D) SVC < 50%, (E) PEF < 75 %, (F) PEF < 50%. Results of ROC analyses for (E) SVC < 80 %, (F) SVC < 50% in the validation cohort. AUC = Area under the curve, cTnT = cardiac Troponin T, NfL = Neurofilament light chain, FVC = Forced vital capacity, SVC = Slow vital capacity, PEF = Peak expiratory flow.

The most significant results were observed for SVC % below 80 % (AUC: 0.75, p = 0.003), FVC below 80% (AUC of = 0.72, p = 0.011), as well as PEF below 75% (AUC of 0.72, p = 0.015).

The optimal cut off-values for serum cTnT to detect impaired respiration measured as FVC, SVC and PEF below 80% and 75 % or 50% range in between 14.00 and 17.80 ng/l, similar to the established cardiac cut-offs (Table 2).

**TABLE 2:**
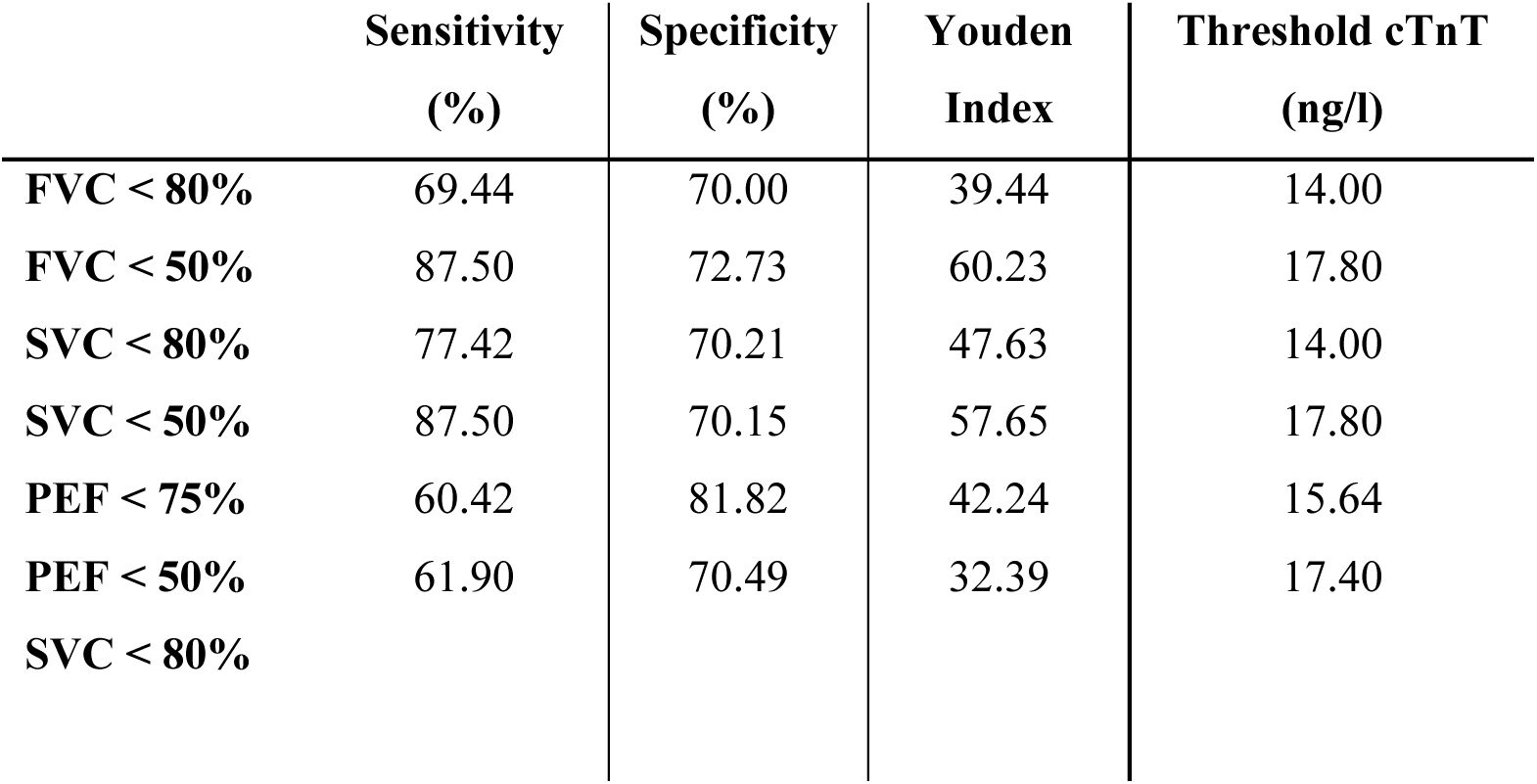
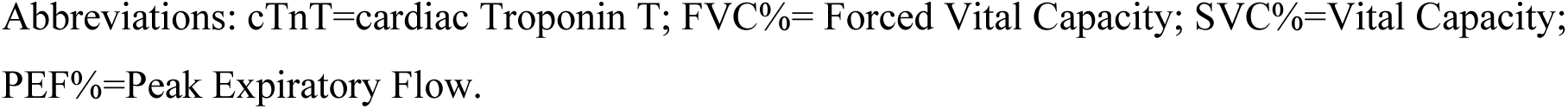
Threshold analysis of cut-offs for serum cTnT to diagnose relevant PFT declines according to Youden index.

### Discussion (909, max:1500)

This is, to the best of our knowledge, the first study reporting the relationship of serum cTnT, or any serum marker for that matter, and pulmonary function tests in patients with ALS. Using a serum marker to obtain insight into respiratory function of a patient might prove immediately useful the everyday clinical practice.

In line with previous literature^6,7,9,10,26,27^, this study confirms that around 65% of ALS patients have elevated serum cTnT levels. The smaller percentage of cTnT elevations in the validation cohort (47%) is explained with the shorter observation time, as serum cTnT tends to raise with disease progression^7^.

The association of serum cTnT with both the ALSFRSr respiratory domain and with the functional PFT parameters, supports the concept that respiratory skeletal muscles drive, or at least substantially contribute to the serum cTnT level elevations in ALS. We note that statistical significance in the validation cohort was not reached for all correlations, but the recapitulation of the key correlation with the ALSFRSr respiratory domain in the smaller cohort (n_v_ =49) underscores the robustness of the core findings (Figure 2).

Information on structural changes in respiratory muscles from ALS patients is scarce.

A single study on PFT parameters and diaphragm atrophy in ALS suggested pre-atrophic muscle changes might have a functional relevance^28^. Supporting this interpretation, our results advocate that cTnT elevations reflect compensatory pre-atrophic remodeling processes in respiratory skeletal muscles. Further research is clearly warranted.

The key finding of our study is the strong inverse correlation between serum cTnT levels with the PFT parameters. Since SVC and FVC, which are measured during full in- and expiration, yield the strongest associations with serum cTnT levels,we propose that the quantity and quality of engaged respiratory muscles drives the serum cTnT level elevation.

Less strong, but still statistically significant associations are found with parameters only involving inspiratory muscle (TLC, IC) or the accessory expiratory muscle groups (FEV1, PEF).

This is in line with the concept that serum cTnT levels might be directly related to the mass of the degenerating respiratory muscle tissue, rather than an individual respiratory muscle group (Figure 3).

Of note, earlier case reports already documented longitudinal measurements of cTnT tracking with progressive respiratory failure and changes in PaO_2_/CO_2_^29,30^. Another study reported that increased cTnT levels were associated with more severe respiratory involvement^24^.

Remarkably, neurofilaments, which have excellent diagnostic and prognostic performance in ALS were *not* correlated with pulmonary function. This striking contrast underscores that serum cTnT captures an aspect of ALS that is missed by neurofilament levels (either in serum or in cerebrospinal fluid). It is well established that neurofilament levels reflect neuroaxonal damage ^31,32^. The published information on the origin of serum cTnT levels in ALS is more circumstantial, but evidence from neuromuscular diseases supports that striated muscle expresses cTnT under pathological conditions^8^.

For the first time we report a serum marker, cTnT, that has good diagnostic accuracy to detect FVC and SVC below the recommended thresholds for NIV indication of 80% and 50%^19,20,34^ and PEF < 75%^35^, associated with limited survival. (Figure 5).

Our calculations provide a rationale for modified cutoff levels of serum cTnT for corresponding pulmonary function impairments. These are similar but not identical to the thresholds that indicate myocardial injury.

Our finding has the potential to significantly expand the biomarker repertoire for ALS. As serum cTnT is a validated biomarker outside the ALS field, many practicalities such as assay standardization, availability and reproducibility are already settled. Future research needs to address how serum cTnT levels respond to therapeutic interventions, e.g. NIV implementation, or investigational compounds in clinical trials^36^.

The data presented here show that cTnT elevations are not just a confounder, but actually allow quantifiable conclusions about the clinical disease course. We believe that this principle can be extended to other neuromuscular diseases where cTnT levels have recently been reported.

### Limitations

A clear limitation of our study is the observational nature of our investigation. We sought to mitigate this by including a discovery and a (albeit small) validation cohort. Also, hospital cohorts, while by nature not rigorously stratified or otherwise standardized, provide robust findings. While the validation cohort does not confirm all correlations found in the discovery cohort, possibly because of lack of power, the overall agreement between the two cohorts strongly argues for the robustness of the observed correlation between cTnT and respiratory function.

Also, PFT assessments in ALS patients are complex and error prone, especially in patients in advanced disease stages (where serum cTnT levels tend to be highest). The confirmation of the findings in the validation cohort strongly mitigates such concerns.

Future, prospective studies should benefit from using more advanced and specific pulmonary measurements, e.g. maximal inspiratory and expiratory pressure.

Although the correlation analyses between serum cTnT and respiratory function parameters yielded statistically significant results, correlations alone cannot establish causality. Further research is clearly necessary to increase the understanding of underlying pathophysiology of the skeletal muscle in ALS patients, and in particular, the respiratory muscles, including the diaphragm.

## Conclusion

We conclude that serum cTnT levels can be repurposed as a biomarker in ALS patients allowing remarkably robust conclusions on the respiratory capacity of ALS patients. Importantly, we identify serum cTnT as marker that is completely independent of neurofilaments. Serum cTnT is emerging as an informative biomarker with diagnostic, categorical and prognostic significance in ALS, substantially complementing the information gained through neurofilament measurements. It will be critical to determine how these two serum markers perform in combination.

## Data Availability

All data produced in the present study are available upon reasonable request to the authors

## Acknowledgements

The research was supported through unrestricted donations from our patients, especially Bruno Schmidt (1965 - 2022) and the Initiative *Alle-Lieben-Schmidt e. V.* to PW.

## Author contribution

T.K, S.B. and P.W. contributed to the conception and design of the study. T.K., P.W., T.G.,F.W.K., L.W., R.F. and S.B. contributed to the acquisition and analysis of data. T.K., S.B., P.W. contributed to drafting the text or preparing figures.

## Data availability

Raw data were generated at the University Hospital Bonn and the Krupp Hospital Essen. Derived data are available from corresponding authors on request.

